# Genome-wide association, Mendelian Randomization and polygenic risk score studies converge on a role of β−amyloid and *APOE* locus in Parkinson disease

**DOI:** 10.1101/2020.07.01.20144493

**Authors:** Laura Ibanez, Jorge A. Bahena, Chengran Yang, Umber Dube, Fabiana G. Farias, John P. Budde, Kristy Bergmann, Carol Brenner-Webster, John C. Morris, Richard J. Perrin, Nigel Cairns, John O’Donnell, Ignacio Álvarez, Monica Diez-Fairen, Miquel Aguilar, Rebecca Miller, Albert A. Davis, Pau Pastor, Paul Kotzbauer, Meghan C. Campbell, Joel S. Perlmutter, Herve Rhinn, Oscar Harari, Carlos Cruchaga, Bruno A. Benitez

**Affiliations:** Department of Psychiatry, Washington University, St. Louis, MO 63110; NeuroGenomics and Informatics. Washington University, St. Louis, MO 63110; Hope Center for Neurologic Disorders, Washington University, St. Louis, MO 63110; Department of Neurology, Washington University, St Louis, MO, 63110; The Charles F. and Joanne Knight Alzheimer Disease Research Center, Washington University School of Medicine, St Louis, MO, 63110; Department of Pathology and Immunology, Washington University, St. Louis, MO 63110; Memory Unit, Department of Neurology, University Hospital Mutua de Terrassa, University of Barcelona, Terrassa, Barcelona, Spain; Fundació per a la Recerca Biomèdica i Social Mútua de Terrassa, University of Barcelona, Terrassa, Barcelona, Spain; Departments of Neuroscience and Radiology, Programs in Physical Therapy and Occupational Therapy, Washington University, St. Louis, MO, 63110; Department of Bioinformatics, Alector, INC, San Francisco, CA, 94080; Department of Genetics, Washington University School of Medicine, St Louis, MO, 63110

**Author notes:** To whom correspondence should be addressed: Bruno A. Benitez, MD, Department of Psychiatry, Washington University School of Medicine, BJC Institute of Health. Box 8134, 425 S. Euclid Ave. St. Louis, Missouri, 63110.

**Keywords:** Parkinson Disease, Genetics, Biomarkers, Alpha-Synuclein, Amyloid Beta, Tau, APOE, polygenic risk scores

## Abstract

Alpha-Synuclein (α-Syn) is the main protein component of Lewy bodies (LB), the pathological hallmark of Parkinson’s disease (PD). Cerebrospinal fluid (CSF) levels of α-Syn are not currently used as a clinical biomarker but may be a proxy for pathological α-Syn accumulation in the brain. Therefore, identifying genetic modifiers of CSF α-Syn levels could provide insights into the underlying molecular mechanisms leading to PD. However, genetic modifiers of CSF α-Syn levels remain unknown. CSF levels of amyloid beta_1-42_ (Aβ42), total tau (t-tau), and phosphorylated tau_181_ (p-tau_181_) are standard biomarkers for the diagnosis of Alzheimer disease (AD); its use as quantitative traits in genetic studies have provided novel insights into AD pathophysiology. A systematic study of the genomic architecture of CSF biomarkers in PD has not been conducted. Here, genome-wide association studies (GWAS) were performed using CSF biomarker levels as quantitative traits in four PD cases and control cohorts (combined N=1,960). CSF biomarker (α-Syn, Aβ42, t-tau, and p-tau_181_) levels were significantly lower in PD cases compared with controls. An SNP, proxy for *APOE* ε4, was associated with CSF Aβ42 levels (effect=-0.5, p=9.2×10^−19^). Several genome-wide suggestive loci associated with CSF α-Syn, t-tau, or p-tau_181_ were found. Polygenic risk scores (PRS) were constructed using the latest PD risk meta-analysis (49,731 PD cases and 784,343 controls) and the largest CSF biomarkers GWAS (N=3,146). PRS calculated using META-PD were associated with PD status in the four cohorts included in the present study (p= 2.2×10^−16^). A highly significant correlation (Nagelkerke pseudo-R^2^ =2.29%; p=2.5×10^−11^) of the genomic architecture between CSF Aβ42 and PD risk was also found. Higher PRS scores were associated with lower CSF Aβ42 levels (p=7.3×10^−04^). Two-sample Mendelian Randomization (MR) approach revealed that CSF Aβ42 plays a role in PD risk (p=1.4×10^−05^) and age at onset (p=7.6×10^−06^), an effect mainly mediated by variants in the *APOE* locus. Subsequently, the *APOE* ε4 allele was associated with significantly lower levels of CSF Aβ42 (p=3.8×10^−06^), higher mean cortical binding potentials (cortical binding of Pittsburgh compound B PET) (p=5.8×10^−08^) and higher Braak Aβ score (p=4.4×10^−04^) in PD participants. Together these results from high-throughput and hypothesis-free approaches (GWAS, PRS and MR) converge on a genetic link between PD with CSF Aβ42 and *APOE*.

## Introduction

Parkinson’s disease (PD) is a neurodegenerative disease characterized by rest tremor, rigidity, bradykinesia, and postural instability.^1^ PD is the most common neurodegenerative movement disorder, affecting more than six million people worldwide, with its prevalence projected to double in the next several decades.^2^ Currently, the diagnosis of PD relies almost entirely on history and clinical examination, as there are no reliable biomarkers for PD. A modest but significant decrease (∼10% to 15%) in cerebrospinal fluid (CSF) alpha-synuclein (α-Syn) levels in PD cases compared with controls^3^ and a correlation with disease progression support the role of CSF α-Syn as a potential PD biomarker.^4-6^ Aggregated and phosphorylated α-Syn is the main protein component of neuronal Lewy bodies (LB) and neurites, the pathological hallmark of Lewy body diseases (LBD). Gene dosage effect of the *SNCA* gene, which encodes α-Syn, correlates with CSF α-Syn levels and a more severe PD phenotype.^7,8^ Additionally, common variants in the *SNCA* promoter are among the top GWAS signals for PD^9^, suggesting that genetic control of CSF α-Syn level plays a role in PD phenotype variability. CSF α-Syn is not currently used as a clinical biomarker^5,10^, but has potential as a proxy for pathological brain α-Syn accumulation.^11^ Therefore, identifying genetic modifiers of CSF α-Syn levels could provide insight into PD pathogenesis. To date, genetic modifiers of CSF α-Syn remain unknown.

The α-Syn accumulation in specific brain regions defines different subtypes of LBD, including neocortical (nLBD); limbic (lLBD); and brainstem or amygdala (bLBD). However, pure α-syn pathology is only found in 45% (bLBD), 32% (lLBD) and 19% (nLBD). Concomitant proteinopathies (presence of Aβ, tau, and TDP-43) are common findings in LBD. Aβ and tau pathology is present in up to 80% and 53% in cases of nLBD, respectively.^12^ LBD with concomitant Alzheimer disease (AD) pathology exhibit a faster cognitive decline.^13^ CSF levels of Aβ42, tau and p-tau are used as proxies of Aβ and tau pathology in the brain.^14^ A correlation between decreased Aβ42 CSF levels and Braak stages of AD neuropathology was found in neuropathologically confirmed LBD.^14^ In cross-sectional studies, PD cases exhibit lower CSF levels of Aβ42 compared to age and gender-matched healthy controls.^15,16^ CSF levels of Aβ42 and total tau (t-tau) change with PD cognitive decline progression.^16^ Decreased CSF Aβ42 levels predict the development of dementia in PD patients.^17,18^ These results suggest that dementia associated CSF biomarker profiles in PD behave similarly to AD. Genome-wide association studies (GWAS) using CSF Aβ42, t-tau, and p-tau_181_ levels as quantitative traits have identified genes involved in AD pathogenesis^19^. However, a systematic study of the role of genetic modifiers of dementia CSF biomarkers in PD has not been thoroughly evaluated yet.

This study aimed to uncover genetic modifiers of α-Syn, Aβ42, t-tau, and p-tau_181_ CSF levels in PD patients by performing a large (N=1,960) GWAS meta-analysis of CSF biomarkers in PD cohorts. PRS and MR analyses were integrated with the latest PD risk meta-GWAS and CSF biomarker summary statistics to examine the causal relationship between CSF biomarkers and PD risk. This is the first comprehensive analysis of CSF biomarkers using GWAS, PRS, and MR in PD.

## Material and Methods

### Study design

The goal of this study was to identify common genetic variants and genes associated with CSF α-Syn, Aβ42, tau, and p-tau_181_ in PD. A three-stage GWAS was used: discovery, replication, and meta-analyses. The discovery phase included 729 individuals from the Protein and Imaging Biomarkers in Parkinson’s disease study (PIB-PD) at the Washington University Movement Disorder Center^20^ (n=103) and the Knight ADRC^19^ (n=626). The replication phase included 1231 independent CSF samples obtained from PD cases and healthy elderly individuals from three additional studies (PPMI, ADNI, and Spain). Meta-analyses were performed using a fixed-effects model. Genetic loci that passed the multiple test correction for GWAS (P<5×10^−8^) were functionally annotated using bioinformatics tools to identify variants and genes driving the GWAS signal. Polygenic risk scores were used to test the correlation between CSF biomarkers and PD risk genetic architecture. Instrumental variables were selected from summary statistics of CSF biomarkers, and Mendelian Randomization methods were applied to test causality with PD risk. PD genetic architecture was obtained from the latest PD risk meta-analysis summary statistics (META-PD).^9^

The Institutional Review Boards of all participating institutions approved the study, and this research was carried out in accordance with the recommended protocols. Written informed consent was obtained from participants or their family members.

### Cohorts/Datasets

This cross-sectional multicenter study was performed using 1,960 samples from non-Hispanic white (NHW) individuals from four cohorts: Washington University in Saint Louis (WUSTL– N=729), the Parkinson’s Progression Markers Initiative (PPMI–N=785), the University Hospital Mutua Terrassa (Spain-N=130) and the Alzheimer Disease Neuroimaging Initiative (ADNI– N=316). Cohorts included 700 clinically diagnosed PD cases, 564 controls, and 386 clinically-diagnosed AD cases. The remaining N=310 individuals do not exhibit symptoms of neurodegenerative disease (Table 1 and Table S1). PD clinical diagnoses were based on the UK Brain Bank criteria.^21^ Clinical, biomarker, and genetic data from the PPMI and ADNI were obtained from the PPMI (www.ppmi-info.org) and ADNI database repository (http://adni.loni.usc.edu/), respectively and accessed most recently on 1 April 2019. Demographic characteristics of some of those cohorts have been published previously.^22-24^ PPMI is a prospective study with ongoing recruitment. CSF samples were obtained at baseline (N=510), six months (N=385), and yearly after enrolment (N_1stYear_=428, N_2ndYear_=404 and N_3rdYear_=320). CSF α-Syn, Aβ42, t-tau, and p-tau_181_ were available for all the mentioned time points in the PPMI cohort.

**Table 1:** Summary Demographics for the individuals with CSF measurements available

|  | **All** | **PD Cases** | **Controls** | **AD Cases** | **Non-neurodegenerative<sup>#</sup>** |
| --- | --- | --- | --- | --- | --- |
| N | 1,960 | 700 | 564 | 386 | 310 |
| Age (years, 95% CI) | 69.3<br>(62.0-75.0) | 66.2<br>(59.6-73.3) | 70.0<br>(64.9-74.3) | 75.0<br>(70.0-80.0) | 64.0<br>(55.0-73.0) |
| Males (N, %) | 1,107<br>(56.5%) | 435<br>(62.1%) | 297<br>(52.7%) | 222<br>(57.5%) | 153<br>(49.4%) |
| Alpha-Synuclein<br>(ZScore(pg/mL)) | -0.02<br>(-0.67-0.65) | -0.03<br>(-0.70-0.60) | 0.02<br>(-0.71-0.71) | 0.14<br>(-0.57-0.80) | -0.23<br>(-0.76-0.47) |
| Amyloid Beta<br>(ZScore(pg/mL)) | -0.20<br>(-0.72-0.64) | -0.20<br>(-0.72-0.45) | 0.11<br>(-0.52-1.04) | -0.63<br>(-1.00- -0.25) | 0.03<br>(-0.49-0.90) |
| Total Tau<br>(ZScore(pg/mL)) | -0.27<br>(-0.66-0.40) | -0.27<br>(-0.70-0.39) | -0.34<br>(-0.67-0.28) | 0.22<br>(-0.41-1.00) | -0.50<br>(-0.73- -0.08) |
| Phosphorylated Tau<br>(ZScore(pg/mL)) | -0.25<br>(-0.69-0.42) | -0.30<br>(-0.72-0.35) | -0.27<br>(-0.70-0.35) | 0.22<br>(-0.42-0.95) | -0.48<br>(-0.81- -0.04) |

\* Concentration values have been standardized using ZScore for comparison purposes. Non-transformed values cannot be compared because there are several measuring methods: SomaScan platform, INNOTEST assay, xMAP-Luminex with INNOBIA AlzBio3, Roche Elecsys cobas e 601 and Euroimmun.
<sup>#</sup> Includes individuals classified as not being AD, PD or having dementia but neither controls such as essential tremor

### Biomarker Measurements

α-Syn in CSF was measured in 107 samples from the WUSTL cohort^20^ and the entire PPMI cohort, using a commercial ELISA kit (Covance, Dedham, MA).^25^ The additional samples (N=622) from WUSTL were quantified using the *SOMAScan* platform (See below). Aβ42, t-tau, and p-tau_181_ were quantified using the INNOTEST assay (WUSTL cohort) and xMAP-Luminex with INNOBIA AlzBio3 (PPMI cohort). The immunoassay platform from Roche Elecsys cobas e 601 was used in the ADNI cohort to quantify all four biomarkers. ELISA assays from Euroimmun (Germany) were used in the Spanish cohort to measure the CSF levels of α-Syn, Aβ42, t-tau, and p-tau_181_. The α-Syn levels were normalized by log_10_ transformation. Aβ42, t-tau, and p-tau_181_ values were normalized by the Z score transformation. Individuals with biomarker levels outside three standard deviations of the mean were removed from the analysis (Table 1).

### PIB Imaging

PIB acquisition and analysis were performed according to published methods.^26^ PET imaging was performed using a Siemens 962 HR ECAT EXACT PET scanner (CTI, Knoxville, TN). Approximately 10-15 mCi of the radiotracer (specific activity ≥ 1,200Ci/mmol) was injected via an antecubital vein, and a 60-minute, three-dimensional (septa retracted) dynamic PET scan was collected in 53 frames (25×5sec, 9×20sec, 10×60sec and 9×300sec). Emission data were corrected for scattering, randoms, attenuation and dead time. Image reconstruction produced images with a final resolution of 6mm full-width half-maximum at the center of the field of view. Frame alignment was corrected for head motion and co-registered to each person’s T1-weighted magnetization-prepared rapid gradient echo (MPRAGE) MR scan.^27^ For quantitative analyses, three-dimensional regions of interest (prefrontal cortex, gyrus rectus, lateral temporal cortex, precuneus, occipital lobe, caudate nucleus, brainstem, and cerebellum) were created by a blinded observer for each subject based on the individual’s MRI scans, with boundaries defined as previously described.^28^ Binding potentials (BP_ND_) were calculated using Logan graphical analysis, with the cerebellum as the reference tissue input function.^28,29^ Mean cortical binding potentials (MCBP) were calculated for each subject as the average of all cortical regions except the occipital lobe. Positron emission tomography (PET) with [11C]-Pittsburgh Compound B (PiB) has identified PD patients with elevated cortical PiB binding corresponding to Aβ plaque deposition at autopsy.^30^ PiB PET is a reliable method for identifying Aβ deposition in patients with Lewy body disorders.^26^

### Neuropathologic analysis

The neuropathological analysis was done at Washington University in Saint Louis, as previously reported.^18^ Briefly, brains were fixed in 10% neutral buffered formalin for two weeks. Paraffin-embedded sections were cut at six μm. Blocks were taken from the frontal, temporal, parietal, and occipital lobes (thalamus, striatum, including the nucleus basalis of Meynert, amygdala, hippocampus, midbrain, pons, medulla oblongata) and the cervical spinal cord. Histologic stains included hematoxylin-eosin and a modified Bielschowsky silver impregnation. Immunohistochemical analysis was performed using the following antibody: Aβ (10D5, Elan Pharmaceuticals). The AD pathologic changes were rated using an amyloid plaque stage (range, 0 to A-C)^31^ and diffuse and neuritic plaques were also assessed, and cases were classified according to the neuropathologic criteria of Khachaturian^32^, the Consortium to Establish a Registry for Alzheimer Disease (CERAD)^33^ and NIA-Reagan.^34^

### Genotyping

All cohorts, except PPMI, were genotyped using the GSA Illumina platform. Genotyping quality control and imputation were performed using SHAPEIT^35^ and IMPUTE2^36^. SNPs with a call rate lower than 98% and autosomal SNPs that were not in Hardy-Weinberg equilibrium (P<1×10^−06^) were excluded from downstream analyses. The X chromosome SNPs were used to determine sex based on heterozygosity rates. Samples in which the genetically inferred sex was discordant with the reported sex were removed. Whole-genome sequence data from the PPMI cohort was merged with imputed genotyping data; only variants present in both files were included in further analysis. Pairwise genome-wide estimates of proportion identity-by-descent tested the presence of unexpected duplicates and cryptically related samples (Pihat>0.30). Unexpected duplicates were removed; the sample with a higher genotyping rate in the merged file was kept for those cryptically related samples. Finally, principal components were calculated using HapMap as an anchor. Only samples with European descent, an overall call rate higher than 95%, and variants with minor allele frequency (MAF) greater than 5% were included in the analyses.

### Single Variant Analysis

The three-stage single variant analysis was performed due to differences in time and platform for biomarker quantification. PLINK1.9^37,38^ was used to perform the analysis of each cohort independently. A linear model corrected by sex, age, and the first two principal components was used. Results from WUSTL, PPMI, ADNI, and Spanish cohort analyses were meta-analyzed using METAL.^39^ For the α-Syn analyses, the WUSTL cohort was divided into two samples quantified using ELISA based method and samples quantified using the SomaScan platform.

### Analysis of Variance

To test the amount of variance explained by specific loci, the genome-wide complex traits analysis (GCTA)^40^ software was used. GCTA estimates the degree to which the phenotypic variance is explained by the provided variants for a complex trait by fitting the effect of these SNPs as random effects in a linear mixed model. It relies on substantial and homogeneous populations.

### Multi-Tissue Analysis

The levels of α-Syn were measured in CSF, plasma, and brain (parietal cortex) using an aptamer-based approach (SomaScan platform). After stringent quality control, CSF (n=835), plasma (n=529), and brain (n=380) samples were included in the downstream analyses (Table S2). The protein level was 10-based log-transformed to approximate the normal distribution and used as phenotype for the subsequent GWAS. The single variant analysis was used in each tissue independently using PLINK1.9^37^. A multi-tissue analysis using the multi-trait analysis of GWAS (MTAG)^41^ was performed to increase the power of detecting a no tissue-specific protein quantitative trait loci (pQTL) for α-Syn. MTAG calculates the trait-specific effect estimate for each tissue separately and then performs the meta-analysis while accounting for possible sample overlap. Measurements of Aβ42, t-tau, and p-tau_181_ were not available in different tissues.

### Polygenic Risk Score (PRS)

The PRSice2 software^42,43^ was used to test if the genetic architecture of PD risk (using the summary statistics from the last PD meta-analysis^9^) was correlated with that of α-Syn CSF levels or with the levels of the other biomarkers in CSF (Aβ42, t-tau, and p-tau_181_) (omics.wustl.edu).^44^ Briefly, the approach constructs a PRS by summing all trait-associated alleles in a target sample, weighted by the effect size of each allele in a base GWAS. SNPs in linkage disequilibrium (LD) are grouped together to avoid extra weight into a single marker. PRSs are computed at various p-value thresholds using the base GWAS. The optimal threshold is considered the one that explains the maximum variance in the target sample. The association was tested using the default parameters and nine p-value cutoffs: 5.0×10^−08^, 5.0×10^−06^, 1.0×10^−05^, 1.0×10^−03^, 0.01, 0.05, 0.2, 0.5 and 1. Longitudinal measures of CSF α-Syn, Aβ42, t-tau, and p-tau_181_ were available for the PPMI cohort. A simplified PRS (detailed below) was used to test if the genetic architecture of PD risk was predictive of biomarker level progression by means of a mixed model. The PD risk PRS using sentinel SNPs from the PD meta-analysis^9^ was modeled using the method described by the Psychiatric Consortium^45^ and ourselves.^46-48^ Briefly, only genetic variants corresponding to the top hit on each GWAS locus (also known as sentinel SNP) available in the dataset with a minimum call rate of 85% were included in the PRS. If not possible, a proxy with R^2^>0.90 was used. The weight of each variant was calculated using the binary logarithm transformation of the reported Odd ratios (ORs). The final PRS is the sum of the weighted values for the alternate allele of all the sentinel SNPs.^46,47^

### Mendelian Randomization

The latest meta-GWAS of PD genetic risk was used^9^. Briefly, Nalls et al. performed a fixed-effects meta-analysis of 17 datasets from European ancestry individuals, analyzing 7.8 million SNPs in 37,688 diagnosed cases, 18,618 UK Biobank proxy-cases (individuals who do not have PD but have a first-degree relative who does), and 1.4 million controls. Summary statistics from the largest GWAS of CSF Aβ42, t-tau, and p-tau_181_ were also used.^44^ Deming et al. performed a one-stage GWAS for 3,146 individuals of European ancestry across nine independent studies.^44^ None of these cohorts included PD affected individuals for each biomarker (Aβ42, t-tau, and p-tau_181_). Finally, the summary statistics of α-Syn CSF levels generated in the current study were used. There was no overlap between CSF biomarker datasets and PD risk datasets.

Two-sample Mendelian randomization to estimate causal effects using the Wald ratio for single variants and an inverse-variance–weighted (IVW) fixed-effects meta-analysis for an overall estimate^49^ was performed using the R package “MendelianRandomization”^50^ (version 0.4.1). To account for potential violations of the assumptions underlying the IVW analysis, a sensitivity analysis using MR-Egger regression and the weighted median estimator^49^ were conducted. Robust regression to downplay the contribution to the causal estimate of instrumental variables with heterogeneous ratio estimates were also performed.^51,52^ Instrumental variables for each GWAS were obtained by clumping each GWAS summary statistics based on a significance threshold of 1×10^−5^ using PLINK1.9.^37^ Instrumental variables were restricted to those that are uncorrelated (in linkage equilibrium) by setting the --clump-r2 flag to 0.0 and the --clump-kb flag to 1000 (1 Mb). Heterogeneity (i.e., instrument strength) was tested using the I^2^ statistic.^53^

## Results

### Association of CSF biomarkers with disease status

A generalized linear model (CSF biomarker levels∼Age+Sex+PD_Status_) including PD cases (N=700) and controls (N=189) from two independent datasets (WUSTL and PPMI –Table S1) in which α-Syn levels were measured with the same platform revealed that all CSF biomarker levels were significantly lower in PD cases compared with healthy-controls (α-Syn: beta_PD_=-0.05, p=2.10×10^−04^; Aβ42: beta_PD_=-0.34, p=4.38×10^−05^; t-tau: beta_PD_=-0.23, p=4.58×10^−03^; and p-tau_181_: beta_PD_=-0.25, p=2.46×10^−03^ – Figure 1). All associations passed multiple test correction (p<0.013). Using a longitudinal model adjusted by age at lumbar puncture, sex, and the first two principal components, significant changes over time were found for CSF Aβ42 (p=0.01) but not for α-Syn, t-tau, or p-tau_181_ in the PPMI cohort (N=785). These results demonstrate that CSF AD biomarkers are associated with PD status.

**Figure 1.**
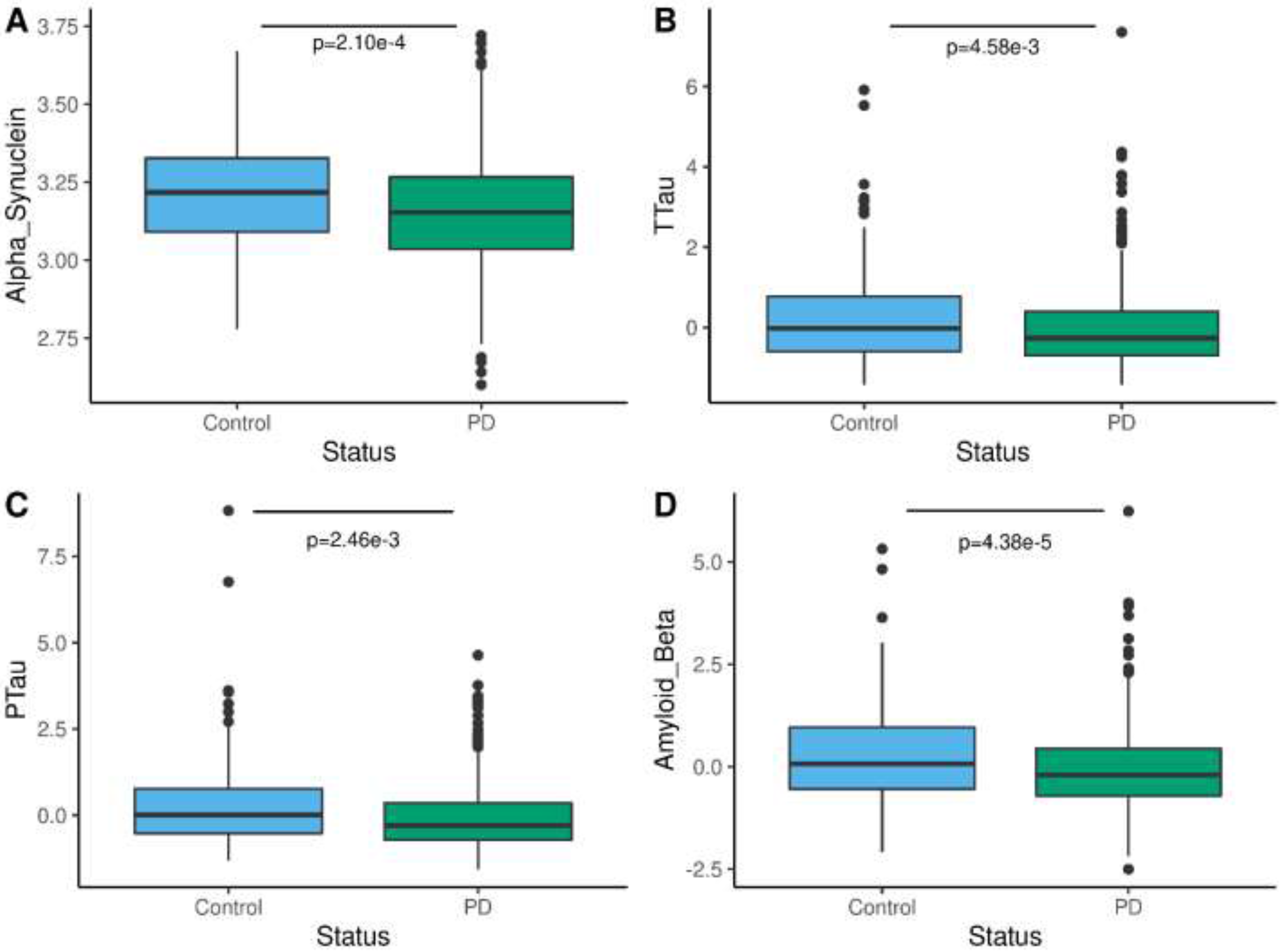
CSF α-Syn, Aβ42, t-tau, and p-tau_181_ levels are lower in PD than in controls. Box plot of the normalized CSF levels of **A**. α-Syn. **B**. total tau. **C**. phosphorylated tau and **D**. Aβ42 in controls (gray) and PD cases (orange). PD cases (N=700) and controls (N=189) from two independent datasets (WUSTL and PPMI). The means for each group are represented by a horizontal line. A generalized linear model (CSF biomarker levels∼Age+Sex+PD_Status_) was used to calculate the statistical differences between the CSF protein levels in PD cases and controls.

### No significant loci were identified for CSF α-Syn, t-tau or p-tau_181_ in PD cohorts

Within each cohort, a linear regression testing the additive genetic model of each single nucleotide polymorphism (SNP) for association with CSF protein levels using age, gender, and two principal component factors for population stratification as covariates did not reveal any genome-wide significant loci associated with CSF α-Syn. Although several suggestive loci (p value<10^−6^ to10^−8^) were identified in these analyses (Table S3 and Figure S1), none of them passed a genome-wide significant threshold when cohorts were combined in the meta-analysis (Figure 2A, Table 2, and Table S3).

**Table 2.** Significant and suggestive sentinel variants in the meta-analysis for GWA of CSF α-Syn, Aβ42, t-tau, and p-tau levels.

| SNP | Location | Effect Allele | MAF | Effect | STD Error | P-Value |
| --- | --- | --- | --- | --- | --- | --- |
| **Alpha Synuclein** |  |  |  |  |  |  |
| rs74924741 | 1:86091232 | T | 0.061 | -0.051 | 0.010 | $7.20 \times 10^{-07}$ |
| rs905124 | 3:190657360 | A | 0.360 | 0.023 | 0.005 | $7.22 \times 10^{-06}$ |
| rs76163673 | 15:55519643 | T | 0.082 | 0.045 | 0.009 | $7.85 \times 10^{-07}$ |
| rs4820059 | 22:32346468 | A | 0.366 | 0.023 | 0.005 | $7.60 \times 10^{-06}$ |
| **Total Tau** |  |  |  |  |  |  |
| rs517201 | 1:146528706 | T | 0.238 | 0.170 | 0.038 | $7.40 \times 10^{-06}$ |
| rs57133061 | 2:476531 | A | 0.053 | -0.314 | 0.070 | $8.46 \times 10^{-06}$ |
| rs2055568 | 2:242393574 | A | 0.715 | -0.161 | 0.036 | $5.88 \times 10^{-06}$ |
| rs13066848 | 3:190668204 | T | 0.643 | -0.170 | 0.034 | $4.46 \times 10^{-07}$ |
| rs2101094 | 4:1235643 | A | 0.106 | 0.232 | 0.052 | $8.95 \times 10^{-06}$ |
| rs28656190 | 4:36521271 | A | 0.055 | 0.385 | 0.074 | $1.58 \times 10^{-07}$ |
| rs76987944 | 5:19423843 | T | 0.833 | 0.189 | 0.043 | $8.71 \times 10^{-06}$ |
| rs2453299 | 5:36031870 | A | 0.137 | 0.212 | 0.048 | $8.35 \times 10^{-06}$ |
| rs7830049 | 8:3640771 | A | 0.933 | -0.314 | 0.064 | $8.92 \times 10^{-07}$ |
| rs55803311 | 10:90753135 | A | 0.117 | 0.229 | 0.051 | $7.16 \times 10^{-06}$ |
| rs11232401 | 11:80633016 | T | 0.927 | -0.284 | 0.063 | $6.17 \times 10^{-06}$ |
| rs12888726 | 14:27302413 | A | 0.506 | -0.143 | 0.032 | $9.28 \times 10^{-06}$ |
| rs7160073 | 14:65450630 | A | 0.359 | 0.149 | 0.033 | $8.55 \times 10^{-06}$ |
| rs799103 | 14:82705630 | A | 0.816 | -0.188 | 0.042 | $6.81 \times 10^{-06}$ |
| rs236012 | 16:71305579 | A | 0.235 | 0.180 | 0.039 | $3.14 \times 10^{-06}$ |
| **Phosphorylated Tau** |  |  |  |  |  |  |
| rs517201 | 1:146528706 | T | 0.238 | 0.182 | 0.038 | $1.62 \times 10^{-06}$ |
| rs6659896 | 1:219365421 | T | 0.141 | 0.201 | 0.045 | $9.25 \times 10^{-06}$ |
| rs34911650 | 3:190670658 | C | 0.642 | -0.163 | 0.034 | $1.21 \times 10^{-06}$ |
| rs7686226 | 4:31156405 | A | 0.558 | -0.145 | 0.033 | $9.76 \times 10^{-06}$ |
| rs35487809 | 8:79343275 | A | 0.934 | -0.324 | 0.066 | $1.03 \times 10^{-06}$ |
| rs2502076 | 10:2989837 | T | 0.318 | 0.176 | 0.035 | $6.60 \times 10^{-07}$ |
| rs55803311 | 10:90753135 | A | 0.117 | 0.227 | 0.051 | $8.22 \times 10^{-06}$ |
| rs769449 | 19:45410002 | A | 0.164 | 0.213 | 0.045 | $1.98 \times 10^{-06}$ |
| **Amyloid Beta** |  |  |  |  |  |  |
| rs7368467 | 1:181249490 | A | 0.145 | 0.216 | 0.045 | $1.39 \times 10^{-06}$ |
| rs3920740 | 1:98233221 | A | 0.357 | 0.292 | 0.056 | $1.64 \times 10^{-07}$ |
| rs2708959 | 2:113670415 | A | 0.238 | 0.207 | 0.046 | $5.91 \times 10^{-06}$ |
| rs116413594 | 2:237102380 | A | 0.155 | 0.255 | 0.057 | $8.44 \times 10^{-06}$ |
| rs4993942 | 3:76733503 | T | 0.514 | -0.152 | 0.032 | $2.76 \times 10^{-06}$ |
| rs2174433 | 5:118582779 | C | 0.363 | 0.269 | 0.056 | $1.77 \times 10^{-06}$ |
| rs35978892 | 5:156025784 | A | 0.947 | 0.340 | 0.072 | $2.40 \times 10^{-06}$ |
| rs13172707 | 5:53146524 | T | 0.210 | 0.175 | 0.039 | $6.66 \times 10^{-06}$ |
| rs3752417 | 6:26045905 | C | 0.230 | 0.212 | 0.043 | $1.02 \times 10^{-06}$ |
| rs2855458 | 6:33154698 | C | 0.239 | 0.231 | 0.042 | $2.88 \times 10^{-08}$ |
| rs17435959 | 11:102651313 | C | 0.276 | 0.243 | 0.050 | $1.11 \times 10^{-06}$ |
| rs4466925 | 12:34380058 | T | 0.167 | -0.195 | 0.043 | $5.95 \times 10^{-06}$ |
| rs769449 | 19:45410002 | A | 0.168 | -0.570 | 0.041 | $4.46 \times 10^{-43}$ |

**Figure 2.**
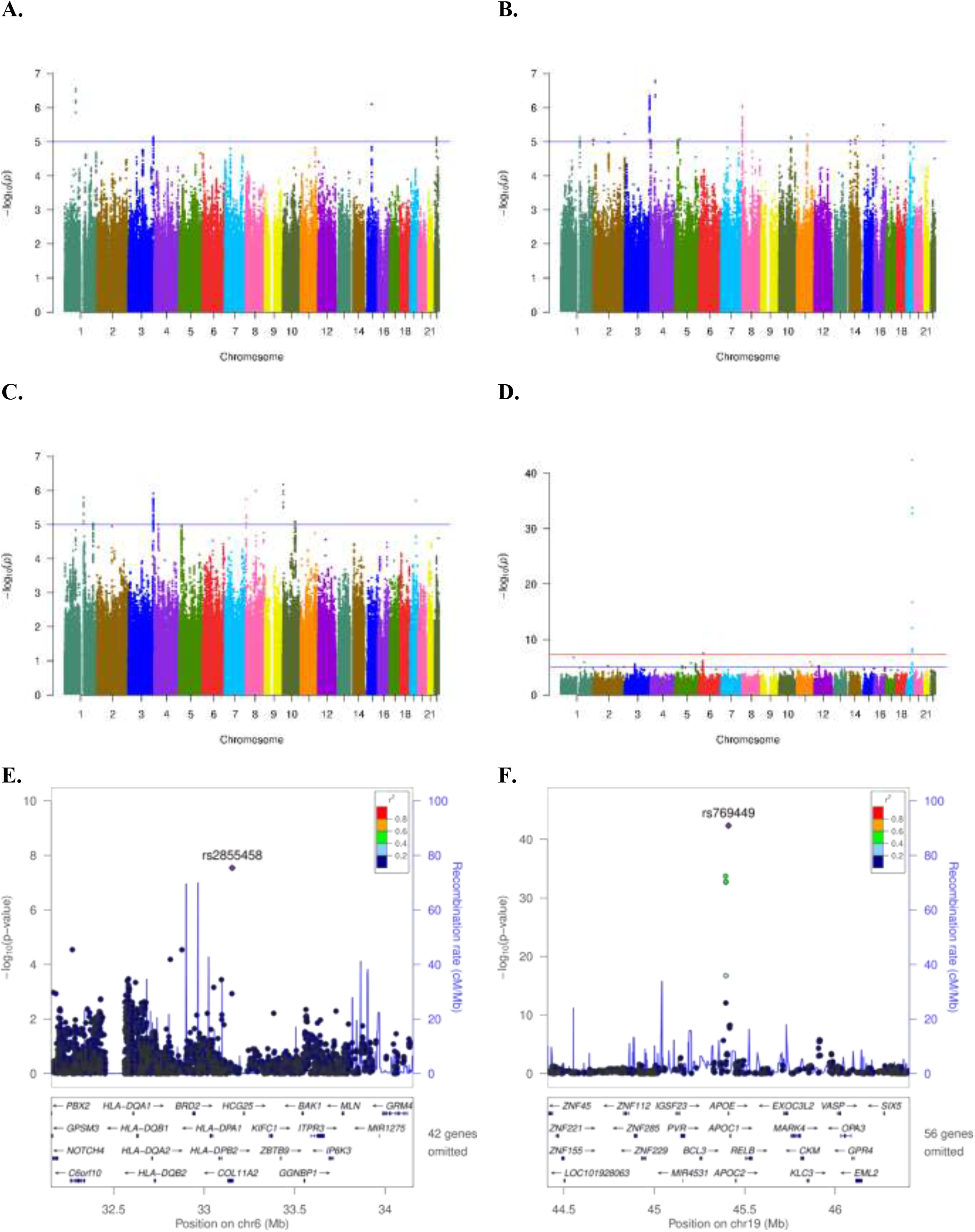
Association plot of single variant analyses of CSF α-Syn, t-tau, p-tau_181_ and Aβ42 levels. Manhattan plot shows negative log_10_-transformed p-values from the meta-analysis of **A. α**-Syn. **B**. total tau. **C**. phosphorylated tau and **D**. Aβ42 CSF levels. The lowest p-value on chr19 (*APOE* locus) was p=4.5×10^−43^. The horizontal lines represent the genome-wide significance threshold, p=5×10^−8^ (red) and suggestive threshold, p=1×10^−5^ (blue). **E, F** Regional association plots of loci are shown for SNPs associated with CSF Aβ42 levels near *HLA* (**E**) and near *APOE* locus (**F**). The SNPs labeled on each regional plot had the lowest p-value at each locus and are represented by a purple diamond. Each dot represents an SNP, and dot colors indicate linkage disequilibrium with the labeled SNP. Blue vertical lines show the recombination rate marked on the right-hand y-axis of each regional plot. Suggestive SNPs for α-Syn, t-tau, p-tau_181_ can be found in Tables S3 to S6.

Joint analysis for CSF α-Syn levels stratifying by PD cases (N=700), PD cases and controls (N=889), AD cases only (N=386), AD cases and controls (N=575) and controls only (N=189) were also performed. None of these analyses revealed any genome-wide significant locus, suggesting that these sample sizes might be underpowered to uncover the genetic modifiers of CSF α-Syn.

For t-tau, individual cohort analyses revealed four genome-wide significant loci and more than 20 loci that were suggestive (Table S5 and Figure S3). However, none of them remained significant in the fixed effect meta-analyses (Figure 2B, Tables 2 and S5). For p-tau_181_, individual cohort analyses revealed three genome-wide significant loci for p-tau_181_ and several suggestive signals (Table S6 and Figure S4). However, none achieved significance in the fixed effect meta-analyses (Figure 1C, Tables 2 and S6).

### Multi-tissue α-Syn genetic GWAS analyses

In a subgroup of samples, α-Syn levels were measured in plasma (N=529), brain (N=380), and CSF (N=835) using the SomaScan platform (Table S2). Single variant analysis was performed in each tissue separately (Figure S2A to S2C). The multi-tissue analysis was performed using MTAG.^41^ This analysis increases the statistical power of a GWAS by increasing the sample size and removing the variance specific to each tissue, thereby increasing the chances of finding genetic modifiers for α-Syn. Although some suggestive loci were observed in chromosomes 3 and 13 (Table S4 and Figure S2D) within genomic regions enriched with long intergenic non-protein coding (LINC) genes (Figure S2E and F), no genome-wide significant locus was identified. These results suggest that the power boost of using MTAG is not enough to unveil the genetic architecture of α-Syn.

### *APOE* locus is associated with Aβ42 CSF levels in PD cohorts

A proxy SNP for APOE ε4, rs769449, was associated at a genome-wide level with CSF levels of Aβ42 in the WUSTL (effect=-0.56, p=4.15×10^−19^), and ADNI cohorts (effect=-0.73, p=1.25×10^−15^). A suggestive association was found in PPMI (effect=-0.43, p=3.09×10^−07^). There was no association in the Spanish cohort (p>0.05). Additional suggestive loci were also identified (Table S7 and Figure S5). The *APOE* locus (effect=-0.57, p=4.46×10^−43^) and a locus in the *HLA* region (effect=0.23, p=2.88×10^−08^) remained significant in the meta-analysis (Figure 2D to 2F). When the cohorts containing only PD cases and controls were analyzed jointly (WUSTL and PPMI – N=700 cases and 189 controls), the *APOE* locus remained GWAS significant (effect=-0.50, p=9.25×10^−19^); the *HLA* region did not pass multiple test correction (effect=0.22, p=3.58×10^−04^).

In the combined analysis of all cohorts (N=1,960), the *APOE* locus accounted for 36.2% of the CSF Aβ42 levels variance (p=2.35×10^−03^). This estimation should be interpreted with caution due to the sample size. Overall, these results revealed a strong and highly significant association between *APOE* locus and lower CSF Aβ42 levels in PD participants.

### Significant correlation of genomic architecture of PD risk and CSF Aβ42

PRS at different p-value thresholds were used to test if the genetic variants associated with neurodegeneration biomarkers were associated with the genomic architecture of PD risk.^9^ PRS calculated using the latest meta-analysis of PD risk (META-PD) were associated with PD status in the four cohorts included in the present study (p<2.20×10^−16^). No correlation was observed between the genetic architecture of PD risk and that of CSF α-Syn, t-tau, or p-tau_181_ (Figure 3).

**Figure 3.**
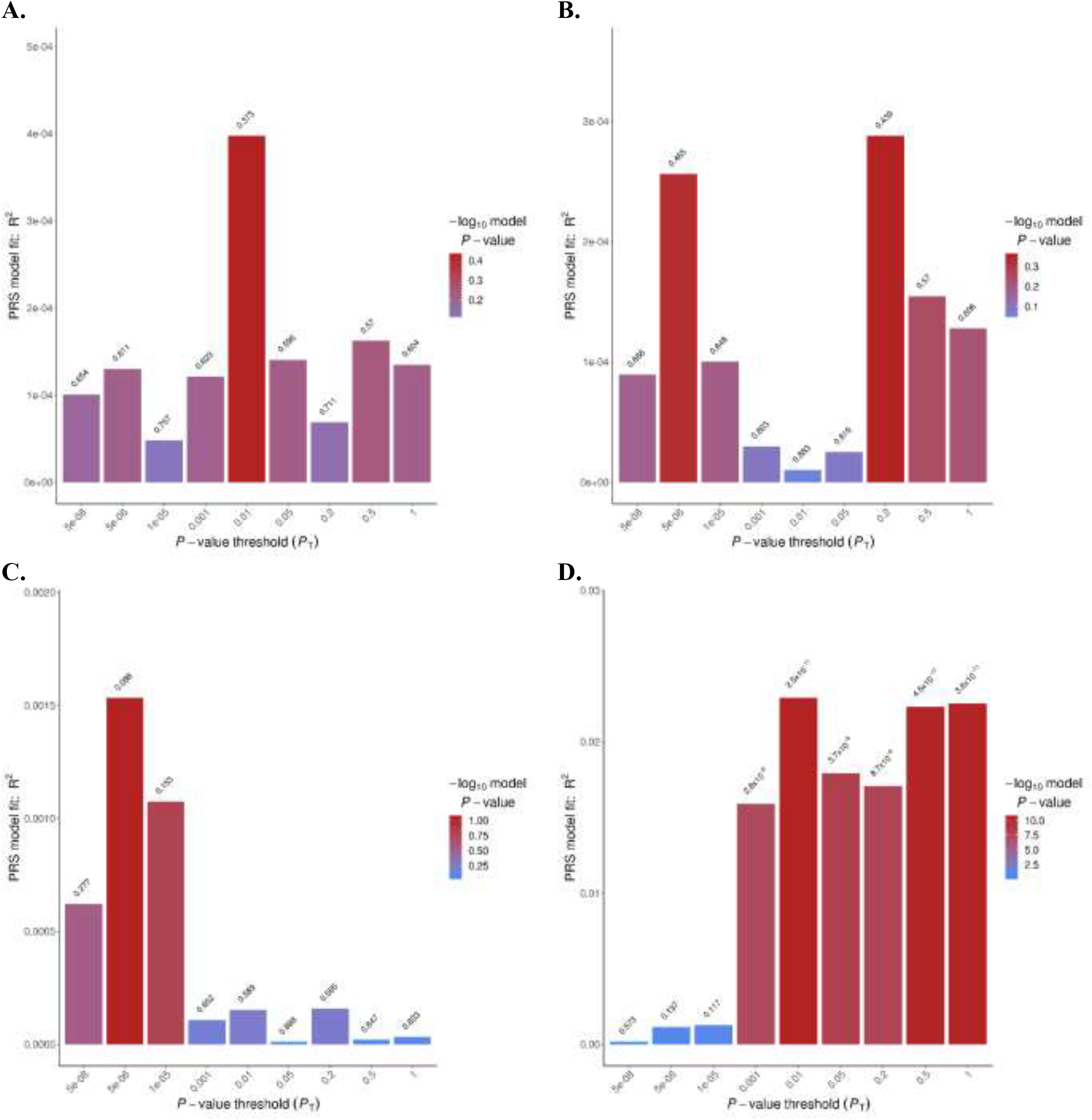
Genetic architecture correlations of PD risk with CSF α-Syn, t-tau, p-tau_181_ and Aβ42 levels. PRSice sum sum bar plots for PD risk and CSF biomarkers. Nagelkerke pseudo-R-squared fit for the model of **A**. CSF α-Syn levels polygenic risk scores (PRS) and PD risk. **B**. CSF t-tau levels PRS—PD risk. **C**. CSF p-tau_181_ PRS and PD risk. **D**. CSF Aβ42 levels PRS and PD risk. Total variance explained by the PRS for multiple p-value thresholds for the inclusion of SNPs, with the red bar indicating the optimal p-value threshold (P_T_), explaining the maximum amount of variance (R^2^) in PD risk in the target sample.

In contrast, the genetic architecture of Aβ42 was correlated with PD risk, with the best fit when collapsing independent SNPs with p-value<0.01 (p=2.50×10^−11^) with a correlation coefficient (R^2^) of 2.29%. The correlation remained significant (p=4.78×10^−08^) with an R^2^ of 2.36% after collapsing independent SNPs with p value<0.01 in PD cases and controls only. In PD patients with both GWAS and CSF biomarker data, the CSF levels of each biomarker was analyzed by quartiles of the PRS calculated from META-PD risk. A significant difference (p=7.30×10^−04^) was found among the top and the bottom quartiles; higher PRS values exhibit lower levels of CSF Aβ42 (Figure S6). No association between PD PRS and longitudinal changes of α-Syn, Aβ42, t-tau, and p-tau_181_ was found. These results indicate that PD risk and Aβ42 CSF levels share genomic architecture.

### Mendelian Randomization suggest a link between CSF Aβ42 and PD

Robust regression with the MR-Egger method found no association for t-tau (effect=-0.33; p=0.25) or p-tau_181_ (effect=-0.10; p=0.79), but revealed a trend towards significance for α-Syn (effect=-1.40; p=0.06), and a significant causal effect for Aβ42 on PD risk (effect=0.43; p=1.44×10^−05^) (Table 3 and S8; Figure 4A to 4C and S7). When each cohort included in the PD meta-analysis was tested separately, Aβ42 showed a causal effect in Nalls 2014^54^ and Nalls 2019^9^ (p=1.54×10^−07^ and 8.74×10^−05^ respectively), but not in Chang 2017^55^ (Table S8). Additionally, a significant causal effect for Aβ42 on PD age-at-onset was found using the data from Blauwendraat, 2019^56^ (effect=7.75; p=7.65×10^−06^ – Table S8). A leave-one-out sensitivity analysis on Aβ42 revealed that the proxy SNP for *APOE* ε4, rs769449 is the main driver of the causal effect of Aβ42 on PD risk. Other SNPs contribute in a smaller proportion to the causal effect (Figure 4D). The MR analysis removing the rs769449 decreased the I^2^ statistic (I^2^=0.00%) and increased the p-value to not-significant levels while removing any of the other variants maintained a high I^2^ (I^2^>95%) and a significant p-value. Altogether these results suggest a causal role of variants on the *APOE* locus and CSF Aβ42 on PD. All the MR methods for α-Syn, t-tau and p-tau_181_ and Aβ42 did not produce significant results when using the META-PD summary statistics or de individual datasets (Table S8 and Figure S7).

**Table 3.** Mendelian randomization results for the causal role of α-Syn, Aβ42, tau, and t-tau in PD using the robust regression MR-Egger method with robust regression

| **Biomarker** | **PD Risk<sup>9</sup>** |  | **PD Age at Onset<sup>56</sup>** |  |
| --- | --- | --- | --- | --- |
|  | **Effect** | **P-Value** | **Effect** | **P-Value** |
| Alpha-Synuclein | -1.389 | 0.064 | -11.018 | 0.835 |
| Amyloid-Beta | 0.430 | 1.44x10<sup>-05</sup> | 7.746 | 7.65x10<sup>-06</sup> |
| Total Tau | -0.338 | 0.246 | 11.276 | 0.069 |
| Phosphorylated Tau | -0.096 | 0.785 | -0.298 | 0.912 |

**Figure 4.**
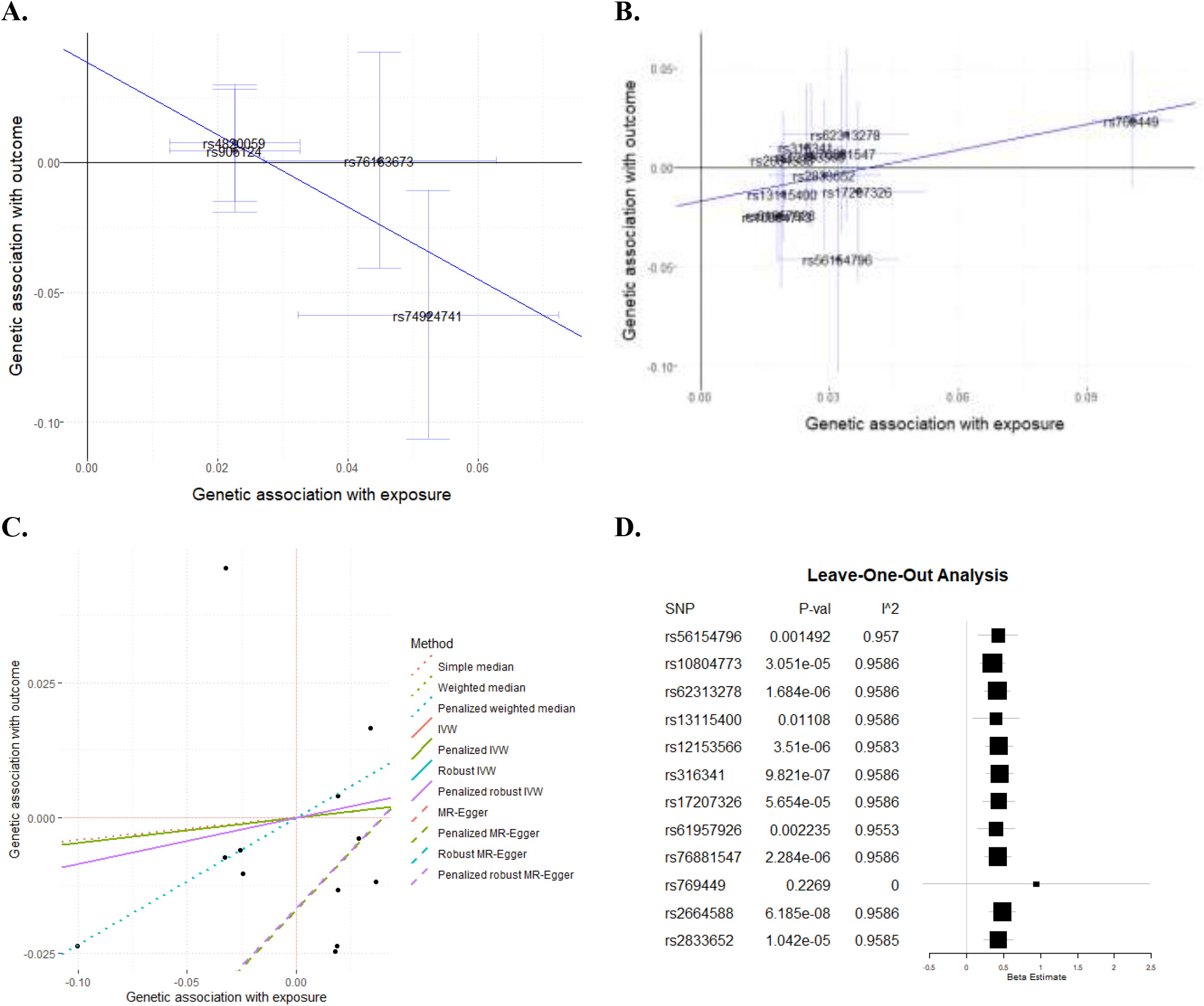
MR regressions on PD risk genetic architecture and CSF α-Syn and Aβ42 levels. **A**. Association between META-PD risk and CSF α-Syn levels (four variants). Robust regression MR-Egger method effect=-1.40 and p=0.06, which is not consistent with causality. **B**. Association between META-PD risk and CSF Aβ42 levels (twelve variants). Robust regression with MR-Egger method effect=0.43 and p=1.44×10^−05^, which is consistent with causality. Each dot corresponds to one genetic variant, with a 95% confidence interval (CI) of its genetic association with the exposure (α-Syn and Aβ42 levels) and the outcome (META-PD risk). Regression lines correspond to the robust MR-Egger method regression; numerical results are given for all tested methods in Table S8. **C**. CSF Aβ42 regression using multiple MR methods. Each dot is one of the twelve variants included in this test; the effect of CSF Aβ42 levels on the x-axis and PD risk on the y-axis. Each line represents the regression of one MR-method of CSF Aβ42 levels on PD risk with one MR method. Additional details on the data sources and analysis methods to generate these figures are provided in Table S8. **D**. The forest plot illustrates the leave-one-out sensitivity analysis between CSF Aβ42 and META-PD risk. MR analysis without rs769449 decreased the I^2^ statistic (I^2^=0.0%) and increased the p-value to non-significant levels, suggesting that the association is mainly driven by this variant.

### *APOE* ε4 is associated with Aβ deposition in brains of PD individuals

CSF Aβ42 and *APOE* genotype data were available for a total of 134 participants, including healthy controls (N=26) and PD participants (N=108). No difference in the *APOE* ε4 frequency was found between PD cases (0.14%) compared to controls (0.11%). However, the CSF Aβ42 levels were significantly different between controls (p=3.00×10^−02^) and PD cases (p=3.80×10^−06^) when stratifying by the presence of *APOE* ε4 allele (Figure 5A), as we have previously reported with a smaller sample size.^20^

**Figure 5.**
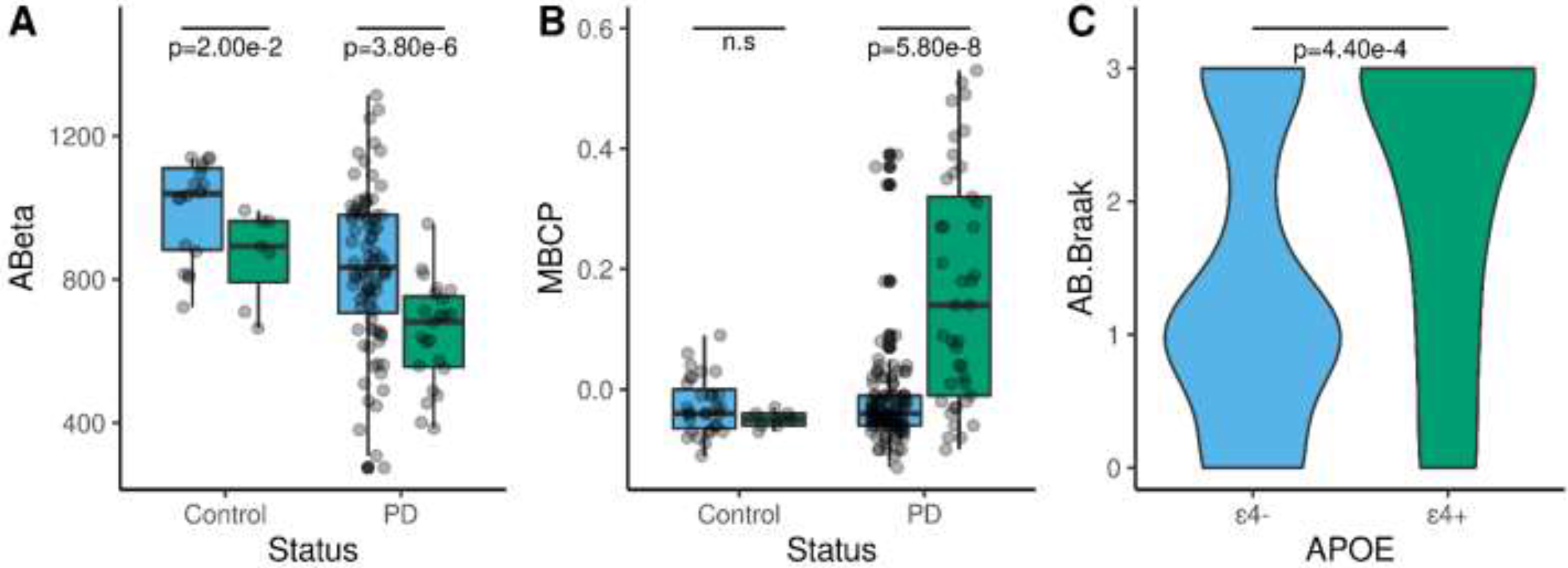
*APOE* ε4 is associated with Aβ42 deposition in the brains of PD individuals. **A**. Comparison of the levels of CSF Aβ42 in control (N=26) and PD (N=108) participants stratified by the presence (ε4+; green) or absence (ε4-; blue) of the *APOE* ε4 allele. **B**. Effect of *APOE* ε4 allele on the levels of mean cortical binding potentials (MCBP) in controls (N=44) and PD (N=156). **C**. PD patients carrying the *APOE* ε4 allele exhibit a higher Braak Aβ score than non-carriers (N=92). Differences between *APOE* ε4 carriers and non-carriers were statistically significant by the Mann-Whitney U test.

PET PiB analysis revealed a MCBP that increased with age-at-onset (r=0.20, p=3.00×10^−02^) and number of *APOE* ε4 alleles (r=0.22, p=8.00×10^−03^) (Figure 5A), but decreased CSF Aβ42 (r=-0.55, p=3.33×10^−12^) in 108 PD patients. A linear regression model, indicates that CSF Aβ42 and *APOE* ε4, explain 48% of the variance of MCBP. *APOE* ε4 is also significantly associated with MCBP (β=0.14, p=1.40×10^−06^) in an analysis that included 200 participants including sex and age as covariates. *APOE* ε4 and age at onset (AAO) explain 20 % of the MCBP variance in this larger cohort. The presence of *APOE* ε4 did not affect the MCBP in healthy controls (p=0.19). However, PD patients carrying *APOE* ε4 exhibit significantly (p=5.80×10^−08^) higher levels of MCBP than non-carriers (Figure 5B).

Neuropathological data and *APOE* genotype were available from 92 PD cases. Individuals carrying an *APOE* ε4 allele had significantly (p=4.40×10^−04^) higher Braak Aβ stage (Figure 5C). *APOE* ε4 correlated with Braak Aβ stage (r=0.33, p=1.00×10^−03^) and diffuse plaques (r=0.42, p=5.00×10^−03^), but not with neuritic plaques (r=0.42, p=0.12). The best multiple linear regression model for the Braak Aβ stage, which included AAO and *APOE* ε4, explained 42% of the variance of the Braak Aβ stage. All these results suggest that *APOE* ε4 drives the Aβ deposition in PD participants.

## Discussion

Here, CSF α-Syn, Aβ42, t-tau, and p-tau_181_ levels were significantly lower in PD cases compared with controls, as we have previously reported with smaller sample size.^20^ GWAS were performed using CSF biomarker levels as quantitative traits in a large combined PD cases and healthy controls cohort (N=1,960). Several genome-wide suggestive loci were associated with CSF α-Syn, t-tau, or p-tau_181_ but with the current sample size, no signal was below the GWAS significant threshold. An SNP proxy for *APOE* ε4 was associated with CSF Aβ42 levels in the GWAS. PRS constructed using the latest PD risk (META-PD) and the largest CSF biomarkers GWAS were used to calculate the genetic correlation between PD risk and CSF biomarker level. The PRS calculated using META-PD were associated with PD status in the four cohorts included in the present study and a highly significant correlation of the genomic architecture between CSF Aβ42 and PD risk was also found. Individuals with higher PRS scores exhibit lower CSF Aβ42 levels. Two-sample MR analysis revealed that CSF Aβ42 plays a role in PD risk and age at onset, an effect mainly mediated by variants in the *APOE* locus. Using a subset of participants from the WUSTL cohort with additional clinical and neuropathological data, we found that the *APOE* ε4 allele was associated with significantly lower levels of CSF Aβ42, higher cortical binding of Pittsburgh compound B PET and higher Braak Aβ score.

This is the first comprehensive analysis of CSF α-Syn and AD biomarkers using GWAS, polygenic risk scores, and MR in PD. We found significantly lower levels of CSF α-Syn in PD cases compared to controls in a cross-sectional analysis but no significant differences in the longitudinal study (PPMI). CSF α-syn, as measured with ELISA-based assays, is not a clinically useful diagnostic marker for PD, and utility as an outcome measure for clinical trials or progression is still controversial^5,10^. CSF biomarkers in AD used as quantitative endophenotypes have provided insights into AD pathophysiology.^19^ Here, we used a large CSF α-Syn cohort (N=1,920) to identify genetic modifiers. However, we did not find any genomic locus associated with CSF α-Syn levels. Recently, a GWAS on CSF α-Syn (N=209) reported a genome-wide significant locus^57^ (rs7072338). In the present meta-analyses (N=1,960), the p-value for rs7072338 was not significant (0.99). In the same cohort (ADNI dataset) used by Zhong et al.^57^, we found a nominal association for this SNP (p=0.50×10^−3^). No correlation was found between the genetic architecture of PD risk with cross-sectional or longitudinal CSF α-Syn levels, consistent with what we have previously reported.^47^ Using MR methods, we found a trend towards significance for the association between the CSF α-Syn levels and the risk of developing PD. However, sensitivity analyses showed limited power due to the small number of variants included in the analyses.

*MAPT* is one of the most consistently replicated loci associated with PD risk in individuals of European descent.^9^ Accumulation of t-tau and p-tau_181_ is found in PD patients^58^. However, it is not clear if tau accumulation plays a role in PD dementia.^18^ The accumulation of t-tau and p-tau_181_ plays a role in parkinsonism of progressive supranuclear palsy and corticobasal degeneration. However, tau pathology is not generally associated with the movement disorder aspects of idiopathic PD. Our results did not reveal any genome-wide significant loci associated with t-tau or p-tau_181_ CSF levels in PD cohorts. A suggestive signal was found in the same genomic locus (3q26) previously reported for t-tau and p-tau_181_ in AD cohorts.^19^ The same direction but half of the effect size found in PD cohorts compare to previously reported in AD cohorts. In PD cohorts, a suggestive association with the same direction (effect=0.21; p=1.98×10^−6^) was also found between the *APOE* locus and CSF p-tau_181_.^19^ One possibility is that our sample could be underpowered to detect a small effect size. No association was found between CSF t-tau or p-tau_181_ genetic architectures with PD risk. We have previously reported a statistical trend for the association of PD risk with the levels of p-tau_181_.^47^ We did not replicate this trend with the current study with a larger sample size. MR analyses did not support a causal relationship between CSF t-tau or p-tau_181_ and PD.

We found a genome-wide significant association between the *APOE* and CSF Aβ42 levels in two cohorts (WUSTL and ADNI), and the association was borderline significant (p=3.09×10^−07^) in the PPMI cohort, as previously reported.^59^ *APOE* locus remained significant in the meta-analysis that included four cohorts. The association of the *APOE* locus with CSF Aβ42 levels has been previously reported in AD case-control cohorts.^60^ We also found that in PD cohorts, the direction of the effect and the p-value of the association with *APOE* were similar to what was previously reported in AD but with higher effect size (−0.57 in PD compared to −0.10 in AD).^44^ There are differences in the strength of the association across the PD cohorts used in this study. The WUSTL cohort includes PD patients with a longer disease duration than those included in the PPMI cohort and is likely to be enriched in PD patients with cognitive decline.^61^ In contrast, the PPMI cohort includes PD patients recently diagnosed with PD and excludes PD patients with dementia.^61^ The *APOE* locus is the most significant locus associated with sporadic DLB, ^62,63^ PD age at onset^56^ and cognitive decline in PD.^61^ However, *APOE* was not found to be associated with PD in the latest PD risk GWAS.^9^ Here, we found that the genetic architecture of CSF Aβ42 levels was correlated with PD risk. PD patients with higher PRS for PD risk exhibit lower levels of CSF Aβ42, suggesting that similar genes or pathways predispose individuals to brain accumulation of Aβ and risk for developing PD. Low levels of CSF Aβ42 predict cognitive decline in PD.^64^ Further studies, ideally with neuropathological information, stratifying PD individuals by dementia or cognitive status, are needed to clarify the role of *APOE* on PD risk or PD dementia.

MR analyses suggest that Aβ42 could play a causal role in PD. Our MR results consistently identified a causal correlation between the *APOE* locus, CSF Aβ, and PD risk in Nalls, 2014^54^ and Nalls, 2019^9^ but not in Chang, 2017^55^. PD patients and controls were classified and ascertained differently among the cohorts with Nalls, 2014^54^ containing the most clinically-diagnosed PD participants, Chang, 2017^55^ using self-reported PD individuals, and Nalls, 2019^9^ including clinically-diagnosed PD plus UKB-proxy PD participants. Our MR results showed a difference in the strength of the association, which may be due to different criteria for case-control classification. In summary, the results from our high-throughput and hypothesis-free approaches (GWAS, PRS and MR) detected the existence of PD genetic risk linked to CSF Aβ42 and *APOE* locus. Further evidence was provided by showing that PD patients carrying *APOE* ε4 allele presented significantly lower levels of CSF Aβ42 (p=3.8×10^−06^), higher MCBP (PiB) (p=5.80×10^−08^) and higher Braak Aβ score (p=4.40×10^−04^). A synergistic relationship between α-Syn and Aβ pathology has been reported in AD, PD and LBD brains.^65^ The presence of Aβ plaques exacerbates propagation of α-Syn pathology in mouse models.^66^ *APOE* ε4 in AD patients drives the production of Aβ and the accumulation of Aβ fibrils.^67^ *APOE* ε4 markedly exacerbates tau-mediated neurodegeneration in a mouse model of tauopathy.^68^ The role of *APOE* in human synucleinopathies is more complex. In LBD patients, the *APOE* ε4 effect on α-Syn pathology could be dependent on concurrent Aβ and/or tau pathology.^12^ However, *APOE* ε4 also promotes α-Syn pathology independently.^69,70^ We recently showed that *APOE* ε4 aggravated α-Syn phosphorylation, worsened motor impairment, and increased neuroinflammation and neurodegeneration in different mouse models.^71^ The present results suggest a genetic link between *APOE* and CSF Aβ42 and Aβ brain deposition in PD.

### Limitations

This study uses the largest sample size reported to date and yet found no significant genetic modifier of CSF α-Syn levels. However, this study had the power to detect strong genetic associations (effect=0.57, p=4.46×10^−43^), comparable to the association of the *APOE* locus with Aβ CSF values. It is possible that the current sample size is not sufficiently powered to detect signals with a smaller effect. Here, we found lower levels of CSF α-Syn in PD patients, which aligns with previous reports. However, neither PRS nor MR analysis revealed evidence of the causal link of CSF α-Syn with PD risk. It has been reported that α-Syn aggregation is neither necessary nor sufficient for neurodegeneration or clinical parkinsonism.^72,73^. The cohorts used in this study rely on clinical diagnosis rather than neuropathological confirmation, which precludes analyses of correlation between CSF α-Syn levels and pathologic brain accumulation of brain α-Syn. Thus, it is possible that CSF α-Syn levels used here may not be reflecting aggregation in the brain. It is not entirely understood how α-Syn gets into CSF, but its levels in CSF may reflect changes in the balance between α-Syn synthesis, secretion, solubility, or degradation pathways.^10^ Consequently, factors that influence CSF α-Syn level variability remain poorly understood. Factors that may have contributed to the lack of power to replicate or detect genetic modifiers of CSF α-Syn include participant characteristics (PD subtypes, misdiagnosis, comorbidities, medications, disease duration), preanalytical factors (blood contamination at LP), and different assays (measuring various abnormal pathological or normal forms of α-Syn)^74^. A unified and standardized platform, such as α-Syn seeding aggregation assays, is necessary to maximize the power of future genetic analyses of CSF α-Syn levels.^75^

## Conclusions

PD is a heterogeneous disorder with different identifiable clinical-pathological subtypes based on symptom severity and predominance. It is conceivable that more homogeneous PD subtypes could be defined using biomarker-driven, clinical-molecular phenotyping approaches. Our results have implications for designing anti-α-Syn therapeutic trials in which both *APOE* genotype and CSF Aβ42 levels could be highly informative. Markers of AD pathology (CSF Aβ42 and PiB) during life may have important prognostic indications in PD to guide clinical trials for homogeneous patient selection. This study, with 1,960 samples with CSF α-Syn levels, shows that the genomic architecture of α-Syn is complex and not correlated with the genomic landscape of PD risk. Additional studies with larger sample sizes and standardized methods to quantify α-Syn in both CSF and brain are needed to uncover genetic modifiers of α-Syn levels. Our results using high-throughput and hypothesis-free approaches detected the existence of PD genetic risk linked to CSF Aβ42 and *APOE* locus. These findings were further validated by strong significant associations of *APOE* ε4 with Aβ deposition in cortical regions of living and postmortem PD patients.

## Data Availability

All data is available in the Center for Neurogenomics and informatics (NGI) website (https://neurogenomics.wustl.edu/). The summary statistics for all the analyses can be easily explored in the Online Neurodegenerative Trait Integrative Multi-Omics Explorer (ONTIME) (https://omics.wustl.edu) and the Charles F. and Joanne Knight Alzheimer Disease Research Center (https://knightadrc.wustl.edu/research/resourcerequest.htm).

## Acknowledgments

We thank all the participants and their families, as well as the many institutions and their staff.

## Funding

This work was supported by grants from the National Institutes of Health (R01AG044546, P01AG003991, RF1AG053303, R01AG058501, U01AG058922, RF1AG058501, R01AG057777, NS097437, NS075321, NS097799 and NS07532), the Alzheimer Association (NIRG-11-200110, BAND-14-338165, AARG-16-441560, and BFG-15-362540), the American Parkinson Disease Association (APDA), the Greater St. Louis Chapter of the APDA, the Jo Riney Fund, the Barnes Jewish Hospital Foundation (Elliot Stein Family Fund and Parkinson Disease Research Fund), and the Paula and Roger Riney Fund. This work was supported by access to equipment made possible by the Hope Center for Neurological Disorders, and the Departments of Neurology and Psychiatry at Washington University School of Medicine. The recruitment and clinical characterization of research participants at Washington University were supported by NIH P50 AG05681, P01 AG03991, and P01 AG026276.

## Competing Interests

CC receives research support from Biogen, EISAI, Alector, and Parabon. The funders of the study had no role in the collection, analysis, or interpretation of data; in the writing of the report; or in the decision to submit the paper for publication. CC is a member of the advisory board of Vivid genetics, Halia Therapeutics, and ADx Healthcare.

